## Supplementary material for "Modelling the impact of non-pharmaceutical interventions on workplace transmission of SARS-CoV-2 in the home-delivery sector": All supplementary files: SupplementaryTextS1.pdf

### Supplementary Text S1 for "Modelling the impact of non-pharmaceutical interventions on workplace transmission of SARS-CoV-2 in the home-delivery sector": Sector-specific data collected and derivation of parameters

Carl A. Whitfield,[1,2,3] Martie van Tongeren,[4,3] Yang Han,[1] Hua Wei,[4,3] Sarah Daniels,[4,3] Martyn Regan,[5,4,3] David W. Denning,[2,3] Arpana Verma,[4,3] Lorenzo Pellis,[1] University of Manchester COVID-19 Modelling Group,[1] and Ian Hall[1,6,3]

[1]*Department of Mathematics, University of Manchester, Manchester, England*

[2]*Division of Infection, Immunity & Respiratory Medicine,*
*School of Biological Sciences, University of Manchester, Manchester, England*

[3]*Manchester Academic Health Science Centre,*
*University of Manchester, Manchester, England*

[4]*Division of Population Health, Health Services Research & Primary Care,*
*School of Health Sciences, University of Manchester, Manchester, England*

[5]*National COVID-19 Response Centre,*
*UK Health Security Agency, London, England*

[6]*Public Health Advice, Guidance and Expertise,*
*UK Health Security Agency, London, England*

#### S1.1. STAFF AND DELIVERY NUMBERS

To obtain realistic figures for the workplace size and number of consignments, we received data from a parcel delivery company and a logistics company that deliver large items, both in the UK.

From the parcel company, we received company-wide figures for number of consignments (i.e. delivery drops) and number of drivers on-shift from 01/01/2020 to 01/09/2020. First, we fitted a linear generalised additive model (GAM) to the number of consignments, in order to extract a smoothed curve $P^{(p)}(t)$ representing the demand over this period (see figure S1(a)). Next, we used negative binomial regression to extract the weekday dependence for both the number of drivers and the number of parcels. The model means ($\mu$ for drivers and $\lambda$ for deliveries) were parameterised as

$$\mu^{(p)}(t) = \exp\left(\alpha_c^{(p)} + \alpha_p^{(p)} P^{(p)}(t) + \sum_{d=0}^{6} \alpha_d^{(p)} \delta_{d,w(t)}\right) \tag{1}$$

$$\lambda^{(p)}(t) = \exp\left(\beta_c^{(p)} + \beta_p^{(p)} P^{(p)}(t) + \sum_{d=0}^{6} \beta_d^{(p)} \delta_{d,w(t)}\right) \tag{2}$$

$$\text{where } w(t) = (t \mod 7), \tag{3}$$

such that $w(t)$ returns the day of the week (Monday = 0, Tuesday = 1, etc.) and $t = 0$ is a Monday. The weekday dependence for each quantity is shown in S1(b). Finally, we were also provided with weekly data for each site (again for number of consignments and number of drivers). We again fitted a negative binomial regression to this data over the same period, with categorical parameters for each site to account for the differences in demand. The mean of this model was parameterised as

$$\bar{\mu}^{(p)}(u,s) = \exp\left(\bar{\alpha}_c^{(p)} + \bar{\alpha}_p^{(p)} W^{(p)}(u) + \sum_{s'=0}^{N_s-1} \bar{\alpha}_{s'}^{(p)} \delta_{s,s'}\right), \tag{4}$$

$$\bar{\lambda}^{(p)}(u,s) = \exp\left(\bar{\beta}_c^{(p)} + \bar{\beta}_p^{(p)} W^{(p)}(u) + \sum_{s'=0}^{N_s-1} \bar{\beta}_{s'}^{(p)} \delta_{s,s'}\right), \tag{5}$$

$$\tag{6}$$

where $u$ is the time in weeks (e.g w/c 01/01/2020 is $u = 0$), $W^{(p)}(u)$ is the weekly demand (i.e. the total demand $P^{(p)}(t)$ summed over the days in week $u$) and $s$ is the site number (indexed from 0 for all $N_s$ sites).

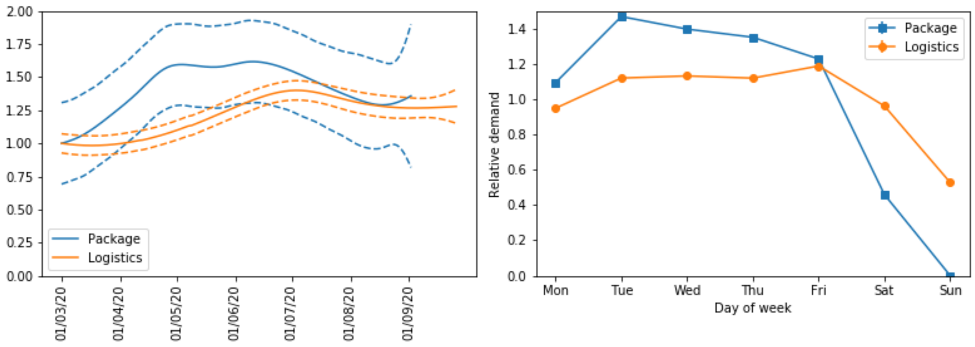


FIG. S1. (a) Smoothed demand curves, fitted using a linear GAM to company-wide figures for number of consignments, for the parcel and logistics companies. The figures are given relative to their value at 01/03/20. (b) Weekday dependence for number of drivers and deliveries fitted using negative binomial regression. Each point shows the number of deliveries or drivers relative to the number on a Friday.

Similar data was provided by a large-items logistics company. In this case, we were provided with daily data for the number of consignments and number of vans for each site in the company from 01/04/19 to 01/09/20. First, we fitted a linear GAM to the total number of consignments over all sites to extract a smoothed demand curve $P^{(l)}(t)$ (shown in S1(a)). Next, we fitted a negative binomial model to both account for weekday effect and site, with mean

$$\mu^{(l)}(t,s) = \exp\left(\alpha_c^{(l)} + \alpha_p^{(l)} P^{(l)}(t) + \sum_{d=0}^{6} \alpha_d^{(l)} \delta_{d,w(t)} + \sum_{s'=0}^{N_s-1} \alpha_{7+s'}^{(l)} \delta_{s,s'}\right). \quad (7)$$

$$\lambda^{(l)}(t,s) = \exp\left(\beta_c^{(l)} + \beta_p^{(l)} P^{(l)}(t) + \sum_{d=0}^{6} \beta_d^{(l)} \delta_{d,w(t)} + \sum_{s'=0}^{N_s-1} \beta_{7+s'}^{(l)} \delta_{s,s'}\right). \quad (8)$$

We assume that, in the absence of COVID isolations, staff numbers on average follow the same daily pattern as driver numbers shown in figure S1(b). The selection of employees in work for a given job $j$ on a given day is a three-step process:

1. All workers in COVID-related isolation are removed.

2. Of those available, each is assigned a random absence with probability $p_{\text{abs}}$ and removed from the availability pool.

3. Of those still available, each has probability $\exp\left(\alpha_{w(t)}^{(i)} - \alpha_{\max}^{(i)}\right)$ of being in work on day $t$, where $\alpha_{\max}^{(i)}$ is the negative binomial parameter corresponding to the day with

highest occupancy. In this notation $i = l$ if in a large-item delivery workplace and $i = p$ is the parcel delivery workplace.

The fitted weekday coefficients are used to model the day-to-day variation in the number of deliveries, so that for $i \in \{l, d\}$

$$D_i(t) = D_i^{(0)}(t) \exp\left[\beta_{w(t)}^{(i)}\right]. \tag{9}$$

We use different values for the background demand $D_i^{(0)}(t)$ based on the type of simulation.

- Scenario simulation: For the parcel delivery workplace, the weekly negative binomial model can be used to approximate the number of deliveries over this period such that

$$D_p^{(0)}(t) = \bar{\lambda}^{(p)}(t, s_{\text{med}}^{(p)}) / \left\{ \sum_{d=0}^{6} \exp\left[\beta_d^{(p)}\right] \right\}, \tag{10}$$

  where $s_{\text{med}}^{(p)}$ is the index of the median site in the parcel delivery dataset. In the large-item delivery workplace then $D_l^{(0)}(t) = \lambda^{(l)}(t, s_{\text{med}}^{(l)})$ where $s_{\text{med}}^{(l)}$ is the index of the median site in the large-item delivery dataset.

- Outbreak simulation: We assume fixed pre-pandemic levels of demand (approximately equal to the scenario simulation case at March 1st 2020). In this case we use $D_p^{(0)}(t) = 3000$ and $D_l^{(0)}(t) = 210$.

in the parcel and large-item workplaces respectively. In this way, $W_{\text{p}}^{(0)}(t)$ represents the number of deliveries that the parcel workplace completes in a week. For outbreak simulations we fix this at

In the case of outbreak simulations, we fix $D_{\text{i}}^{(0)}(t) = N_P$ where $N_P = 3000$ deliveries per week for the parcel company and $N_P = 210$ deliveries per week for the logistics company.

To make the number of staff in work each day consistent with the data for a median workplace, we defined the total number of drivers as

$$N_D = \left\lceil \frac{1}{7} \sum_{w=1}^{7} \frac{N_P}{p_{PD}(1 - p_{\text{abs}})} \exp\left(\beta_{\text{P-DOW}}^{(w(t))} - \beta_{\text{S-DOW}}^{(w(t))} + \beta_{\text{S-DOW}}^{(\max)}\right) \right\rceil, \tag{11}$$

where $p_{PD}$ is the average number of parcels delivered per driver per day under normal conditions (85 for parcel delivery and 15 for large-item). The number of warehouse and office staff were then defined relative to this.

In the in situ scenarios, we use the negative binomial fitted demand such that

$$D_{\rm av}(t) = \exp\left(\beta_c + \beta_p P^{(i)}(t) + \beta_{\rm site}^{(s_{\rm med})}\right) \tag{12}$$

where $\beta_{\rm site}^{(s_{\rm med})}$ is the demand coefficient for the median site and $i \in p, l$ indicates whether it is the parcel or logistics company.

#### S1.2. COMMUNITY INCIDENCE AND PREVALENCE

The estimates community incidence data used for the continuous-source outbreak simulations are shown in figure S2. They are based on data on hospitalisations and deaths from 01/03/2020 – 31/05/2020 inclusive. Community prevalence was simply estimated as the cumulative sum of the incidence from the previous 10 days inclusive.

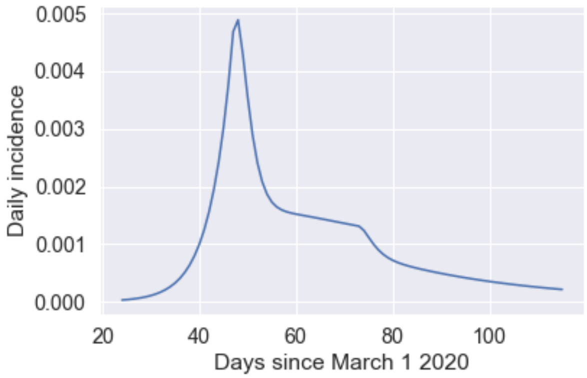


FIG. S2. Community incidence rates assumed for the continuous-source outbreak scenario.

#### S1.3. VIRAL LOAD, INFECTIVIOUSNESS, AND TEST-POSITIVE PROBABILITY

The infectiousness and test-positive probability of individuals over time are generated using the viral-load based model in [1]. The "Ke er al." model of RNA viral-load and infectiousness, as defined in [1] and derived from the data in [2] is used here. The test-sensitivity model for LFD used here is the 'high-sensitivity' model in [1], based on the phase 3b results from Porton Down testing of the Innova LFDs [3]. Sample curves and mean profiles are given in figure S3, reproduced from [1].

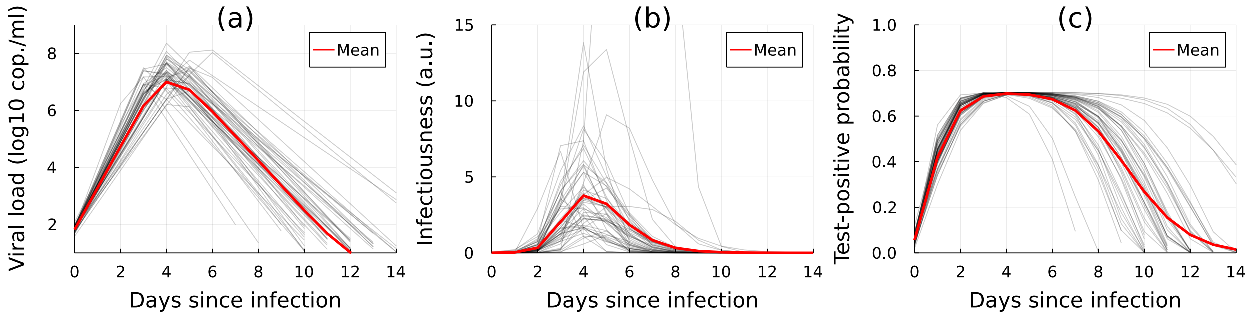


FIG. S3. Each figure shows 50 randomly generated profiles of (a) RNA viral load ($\log_{10}$ copies/ml) and their associated (b) infectiousness (normalised units) and (c) test-positive probability. The red lines show the mean of 10,000 generated individuals at each time point (where a missing value is taken as 0).

#### S1.4. TRANSMISSION RATES

A contact of type $i$ between an infectious individual $k$ and a susceptible person $k'$ is assumed to have transmission rate

$$\gamma_{k,k'}^{(i)} = \beta_{F2F} c_i S_{k'}(t) J_k(t) \tau_i / 2^{x_i - 1} \tag{13}$$

where $\beta_{F2F}$ is the average transmission rate for face-to-face contact while talking at 1m separation, $c_i$ is the transmission modifier for the type of contact, taken from [4], $\tau_i$ is the contact duration, and $x_i$ is the contact distance in metres. $S_{k'}$ and $J_k$ are the susceptibility and infectiousness of individuals $k'$ and $k$ respectively on day $t$.

The baseline transmission rate $\beta_{F2F}$ is estimated using multiple sources. First, in the [4], the probability of transmission while talking at 1m separation for one hour is given as 0.06 for the original SARS-CoV-2 virus and 0.15 for the delta variant. This suggests a $\beta_{F2F}$ of $0.07^{-1}$ or $0.15^{-1}$ respectively. Test and trace data [5] suggests a workplace/school secondary attack rate of $\sim 3\%$ between close contacts, defined as either $< 1$m for more than a minute, 1-2m for $> 15$minutes or sharing a vehicle/skin-to-skin contact. If we take the average contact to be between 1-2m for 15-60min this gives a plausible range of $\beta_{F2F}$ =0.03–0.24h$^{-1}$. Finally, in [6] the probability of infection while talking at 1m separation for 15 mins is $\sim 0.03$, suggesting a $\beta_{F2F}$ of 0.13h$^{-1}$. Therefore, we take $\beta_{F2F} = 0.15$h$^{-1}$ in this model as a "best guess" from this range of estimates.

For household contacts, we use the transmission rate (per day) of

$$\gamma_{k,k'}^{(HH)} = \beta_{HH} S_{k'}(t) J_k(t) \tag{14}$$

where $\beta_{HH} = 0.07$. This was set empirically using $10^6$ realisations of infectiousness trajectories so that the mean probability of household infections was 40% (approximately twice the observed household attack rate from the original SARS-CoV-2 strain [4, 7–9]). Comparing to (13), we see that this means the household transmission rate is approximately equivalent to 30 mins of F2F conversation indoors at a distance of 1m per day.

For package mediated fomites, we use a two-step process such that a fomite infection between workers occurs with probability

$$p_{kk'} = \exp\left[-(t_2 - t_1)\ln 2/\lambda\right]\left[1 - \exp(-\beta_{FOM} J_k(t_1) S_{k'} t_2\right] \tag{15}$$

where $t_1 < t_2$ is the time the package is handled by an infectious individual $k$ and $t_2$ is the time the package is handled by susceptible individual $k'$. The parameter $\lambda$ is the half-life of the viable virus on the packaging, so the first exponential term represents this decay process. The second term is the same as used for all other contacts except that $\beta_{FOM}$ is effectively the probability of transmission if an infected person handles a package that is then immediately handled by another employee.

Package handling is assumed to occur as follows

1. A picker (or picker-pair) $k$ handles all of their $l_k$ packages at times uniformly distributed in a time window of $\tau_L$ hours.

2. Each infectious packages $n$ is then assigned a random integer $r_n$ between 1 and $P$, which then determines which driver (or driver-pair) they get assigned to, such that $1 + \sum_{k'=1}^{k-1} d_k \leq r_n \leq \sum_{k'=1}^{k} d_k$ is assigned to driver $k$.

3. Drivers are assumed to handle all of their $d_k$ packages twice. First they handle all at time $\tau_L$, and second at times distributed between $\tau_L$ and $\tau_L + \tau_D$ (where $\tau_D$ the time is takes to complete all of the deliveries).

Finally, for shared-spaces (not including F2F contact), we use the dose-response relationship originally derived for SARS-CoV [10] and applied to SARS-CoV-2 [11]. Assuming that the number of aerosolised infectious plaque-forming units (PFUs) exhaled per hour is $n_0$, the concentration in the surrounding air (assuming fast mixing) will be

$$c(t) = \frac{n_0}{(\dot{f} + \lambda)V}\left[1 - e^{-(\dot{f}+\lambda)t}\right] \tag{16}$$

where $V$ is the volume of the room and $\dot{f}$ is the air exchange rate (air changes per hour, ACH), and $\lambda = \ln(2)/t_{1/2}$ is the decay rate of the aerosolised virions (such that $t_{1/2}$ is their half-life).

Therefore, the average concentration of infectious PFUs in the room while occupied by a single infectious person for duration $\tau$ is

$$\bar{c}(\tau) = \frac{n_0}{(\dot{f} + \lambda)V}\left[1 - \frac{1}{(\dot{f} + \lambda)\tau}\left(1 - e^{-(\dot{f}+\lambda)\tau}\right)\right] \tag{17}$$

To convert this to a transmission rate, we assume that an average individual has an inhalation rate $\dot{V}_T$ (volume of air inhaled per hour). Their average dose, if sharing the space for the full period $\tau$, is then $\dot{V}_T\bar{c}(\tau)\tau$.

The exponential dose-response model used in [10] has the same form as our infection model, so the transmission rate between individuals sharing the same space for period $\tau$ is given as

$$\beta_{ss} = \frac{\dot{V}_T\bar{c}(\tau)}{ID_e} \tag{18}$$

such that the probability of infection via this route is $p_{ss} = 1 - \exp(-\beta_{ss}\tau)$. The infectious dose $ID_e$ is highly uncertain, as is the dose exhaled $n_0$. However, the ratio $n_0/ID_e$ is approximated in [12] as ∼16–18 per hour at peak infectiousness. Given that, in our model, peak infectiousness is ∼4 times mean infectiousness, we use $n_0/ID_e = 4.0$ here.

In order to more directly compare this to the F2F interactions, we derive an "effective interaction distance" that is implied by this transmission rate. I.e. how far away would the two contacts have to be in a (no-talking) F2F interaction to experience an equivalent transmission rate in our model. This is given by $x_{\text{eff}} = 1 + \log(\beta_{F2F}/5\beta_{ss})$, since $\beta_{F2F}/5$ is the F2F transmission rate at 1m distance with the "no-talking" modifier. The parameters used to estimate $x_{ss}$ are given in table S1.

#### S1.5. MODES OF FACE-TO-FACE CONTACT

We include several routes through which face-to-face contacts occur in the workplace, which are summarised in table 2 (in the main text). Below we give some further details about how these are simulated:

- **Cohort contacts:** Daily F2F interactions between employees working the on the same shift and in the same area. For details of the rationale between how these cohorts are assigned, see Appendix **??**. It is assumed that, each employee contacts all others within their cohort for the same duration, such that the total face-to-face contact time for any given individual in a cohort is fixed. I.e. if a cohort consists of M people, each has a F2F contact with the other $M - 1$ people in their cohort for $\tau_{\text{coh}}/(M - 1)$.

| Parameter | Description | Value | Source |
|---|---|---|---|
| $V$ | Room volume. | 150 $m^3$ | Approximated ($\approx 7 \times 7 \times 3$) |
| $\dot{V}_T$ | Tidal breathing rate. | 0.7 $m^3$ $h^{-1}$ | Approximated |
| $f$ | Air changes per hour. | 2 $h^{-1}$ | Assumed (typical unventilated office value) |
| $\lambda$ | Decay rate of SARS-CoV-2 in air. | 0.76 $h^{-1}$ | [13] |
| $n_0/ID_e$ | Average number of infectious quanta exhaled per hour for an individual infected with SARS-CoV-2 | 4.0 | [12] |

TABLE S1. Parameters used to determine the shared-space transmission parameter $x_{ss}$.

- **Random interactions:** On a given day, each person with job $j$ at work will randomly contact another employee with job $j'$ with probability

$$p_{jj'} = \begin{cases} \rho_D p_c & \text{if } j = D \text{ or } j' = D \\ p_c & \text{otherwise} \end{cases} \tag{19}$$

The factor $\rho_D$ is to account for the reducing mixing in drivers due to the time they spent not at the workplace during a shift. Note, $p_{DD} = \rho_D$ and not $\rho_D^2$ because we are assuming that drivers are on site at approximately the same time as each other.

- Explicit workplace pairings (includes **large-item handling**, **pair delivery**, and **pair delivery** routes): These are necessary for execution of picking and delivery of large items. When these pairings exist, all drivers and pickers are paired at the start of each shift, and this contact is guaranteed to occur during the shift. If the "fixed pairings" intervention is applied, then staff in pairs always have the same partners, otherwise the pairs are assigned randomly at the start of each day.

---

[1] Carl A. Whitfield, University of Manchester COVID-19 Modelling Group, and Ian Hall. Modelling the impact of repeat asymptomatic testing policies for staff on SARS-CoV-2 transmission potential. *arXiv*, October 2022. doi:10.48550/arXiv.2210.08888.

[2] Ruian Ke, Pamela P. Martinez, Rebecca L. Smith, Laura L. Gibson, Agha Mirza, Madison Conte, Nicholas Gallagher, Chun Huai Luo, Junko Jarrett, Ruifeng Zhou, Abigail Conte, Tongyu Liu, Mireille Farjo, Kimberly K. O. Walden, Gloria Rendon, Christopher J. Fields, Leyi Wang, Richard Fredrickson, Darci C. Edmonson, Melinda E. Baughman, Karen K. Chiu, Hannah Choi, Kevin R. Scardina, Shannon Bradley, Stacy L. Gloss, Crystal Reinhart, Jagadeesh Yedetore, Jessica Quicksall, Alyssa N. Owens, John Broach, Bruce Barton, Peter Lazar, William J. Heetderks, Matthew L. Robinson, Heba H. Mostafa, Yukari C. Manabe, Andrew Pekosz, David D. McManus, and Christopher B. Brooke. Daily longitudinal sampling of SARS-CoV-2 infection reveals substantial heterogeneity in infectiousness. *Nat Microbiol*, 7 (5):640–652, May 2022. ISSN 2058-5276. doi:10.1038/s41564-022-01105-z.

[3] Tim Peto. COVID-19: Rapid Antigen detection for SARS-CoV-2 by lateral flow assay: A national systematic evaluation for mass-testing. *medRxiv*, page 2021.01.13.21249563, 2021.

[9] Qin Long Jing, Ming Jin Liu, Zhou Bin Zhang, Li Qun Fang, Jun Yuan, An Ran Zhang,

Natalie E. Dean, Lei Luo, Meng Meng Ma, Ira Longini, Eben Kenah, Ying Lu, Yu Ma, Neda Jalali, Zhi Cong Yang, and Yang Yang. Household secondary attack rate of COVID-19 and associated determinants in Guangzhou, China: A retrospective cohort study. *Lancet Infect. Dis.*, 20(10):1141–1150, 2020. ISSN 14744457. doi:10.1016/S1473-3099(20)30471-0.

[10] Toru Watanabe, Timothy A. Bartrand, Mark H. Weir, Tatsuo Omura, and Charles N. Haas. Development of a dose-response model for SARS coronavirus. *Risk Anal.*, 30(7):1129–1138, 2010. ISSN 02724332. doi:10.1111/j.1539-6924.2010.01427.x.

[11] Xiaole Zhang and Jing Wang. Dose-response Relation Deduced for Coronaviruses from COVID-19, SARS and MERS Meta-analysis Results and its Application for Infection Risk Assessment of Aerosol Transmission. *Clin. Infect. Dis.*, 2020. ISSN 1058-4838. doi: 10.1093/cid/ciaa1675.

[12] Florian Poydenot, Ismael Abdourahamane, Elsa Caplain, Samuel Der, Jacques Haiech, Antoine Jallon, Inés Khoutami, Amir Loucif, Emil Marinov, and Bruno Andreotti. Risk assessment for long and short range airborne transmission of SARS-CoV-2, indoors and outdoors. *PNAS Nexus*, page pgac223, October 2022. ISSN 2752-6542. doi:10.1093/pnasnexus/pgac223.

[13] Neeltje van Doremalen, Trenton Bushmaker, Dylan H. Morris, Myndi G. Holbrook, Amandine Gamble, Brandi N. Williamson, Azaibi Tamin, Jennifer L. Harcourt, Natalie J. Thornburg, Susan I. Gerber, James O. Lloyd-Smith, Emmie de Wit, and Vincent J. Munster. Aerosol and Surface Stability of SARS-CoV-2 as Compared with SARS-CoV-1. *New England Journal of Medicine*, 382(16):1564–1567, April 2020. ISSN 0028-4793. doi:10.1056/NEJMc2004973.
