## Supplementary material for "Modelling the impact of non-pharmaceutical interventions on workplace transmission of SARS-CoV-2 in the home-delivery sector": All supplementary files: SupplementaryTextS2.pdf

### Supplementary Text S2 for "Modelling the impact of non-pharmaceutical interventions on workplace transmission of SARS-CoV-2 in the home-delivery sector": Simulation Algorithm

Carl A. Whitfield,[1,2,3] Martie van Tongeren,[4,3] Yang Han,[1] Hua Wei,[4,3] Sarah Daniels,[4,3] Martyn Regan,[5,4,3] David W. Denning,[2,3] Arpana Verma,[4,3] Lorenzo Pellis,[1] University of Manchester COVID-19 Modelling Group,[1] and Ian Hall[1,6,3]

[1]*Department of Mathematics, University of Manchester, Manchester, England*

[2]*Division of Infection, Immunity & Respiratory Medicine, School of Biological Sciences, University of Manchester, Manchester, England*

[3]*Manchester Academic Health Science Centre, University of Manchester, Manchester, England*

[4]*Division of Population Health, Health Services Research & Primary Care, School of Health Sciences, University of Manchester, Manchester, England*

[5]*National COVID-19 Response Centre, UK Health Security Agency, London, England*

[6]*Public Health Advice, Guidance and Expertise, UK Health Security Agency, London, England*

At initialisation, the following model features are generated

1. Agents are assigned job roles and susceptible status.
2. The cohort contact network is generated as follows:
   (a) Within each job role, each agent is assigned a cohort at random.
   (b) For each driver cohort, one member of warehouse staff (a 'picker') is assigned to that cohort (representing a supervisory role).
   (c) Within each cohort, all agents are connected by edges with identical weight, which are active only when both agents are in work.
3. The house-share network is generated as follows:
   (a) $NH = \lfloor (N_D + N_L + N_O)/(1 + H) \rceil$ households are generated.
   (b) Each agent is assigned randomly to a household.
   (c) Within each household, all agents are connected by edges with identical weight that are active every day.
4. The car-share network is generated as follows:
   (a) $NC = \lfloor NH/(1 + C) \rceil$ car-shares are generated.
   (b) Each household is assigned to a car-share, with all agents in that household assigned to the same car-share.
   (c) Within each car-share, all agents are connected by edges with identical weight that are active only when both agents are in work.

If simulating an outbreak scenario, an index case is infected on a random start day from Monday to Sunday. Upon infection, an agent is assigned the following:

- Viral load and infectiousness trajectories.
- Symptom onset time.
- Adherence to self-isolation (true with probability $p_{\text{isol}}$).
- If testing: a test positive probability trajectory (see Supplementary Text S3).

The main simulation loop is executed for each day of the simulation, and proceeds as follows:

1. Update infectious state of all individuals moving any to 'Recovered' status who have reached the end of their infectious period.

2. Perform testing (if a testing day). For all positive tests generate an isolation time from the current day as $\lfloor \tau_d + u_{01} \rceil$ where $u_{01} \sim U(0, 1)$ is a number uniformly distributed beteween 0 and 1, and $\lfloor . \rceil$ indicates rounding to the nearest integer.

3. Update isolation status for any who are due to isolate on this day.

4. Randomly generate any new introductions due to community incidence.

5. Select employees in work, all others are excluded from any workplace contacts for the rest of the day.

6. Generate all successful workplace infection events.

7. Generate customer infections and introductions.

8. Remove any infections where the infection target is not in the 'susceptible' state. For any viable targets that are subject to more than one successful infection event, select the recorded infection event at random.

9. Record all infection events, and for every individual infected change their status to 'infected' and their infection time to the current day.

10. Increment the day and return to step 1 unless the maximum number of days has been simulated. If an outbreak simulation and all individuals are in the 'recovered' state, terminate the simulation.
