## Supplementary material for "Modelling the impact of non-pharmaceutical interventions on workplace transmission of SARS-CoV-2 in the home-delivery sector": All supplementary files: SupplementaryTextS3.pdf

### Supplementary Text S3 for "Modelling the impact of non-pharmaceutical interventions on workplace transmission of SARS-CoV-2 in the home-delivery sector": Baseline Modelling Results

Carl A. Whitfield,[1,2,3] Martie van Tongeren,[4,3] Yang Han,[1] Hua Wei,[4,3] Sarah Daniels,[4,3] Martyn Regan,[5,4,3] David W. Denning,[2,3] Arpana Verma,[4,3] Lorenzo Pellis,[1] University of Manchester COVID-19 Modelling Group,[1] and Ian Hall[1,6,3]

[1]*Department of Mathematics, University of Manchester, Manchester, England*

[2]*Division of Infection, Immunity & Respiratory Medicine, School of Biological Sciences, University of Manchester, Manchester, England*

[3]*Manchester Academic Health Science Centre, University of Manchester, Manchester, England*

[4]*Division of Population Health, Health Services Research & Primary Care, School of Health Sciences, University of Manchester, Manchester, England*

[5]*National COVID-19 Response Centre, UK Health Security Agency, London, England*

[6]*Public Health Advice, Guidance and Expertise, UK Health Security Agency, London, England*

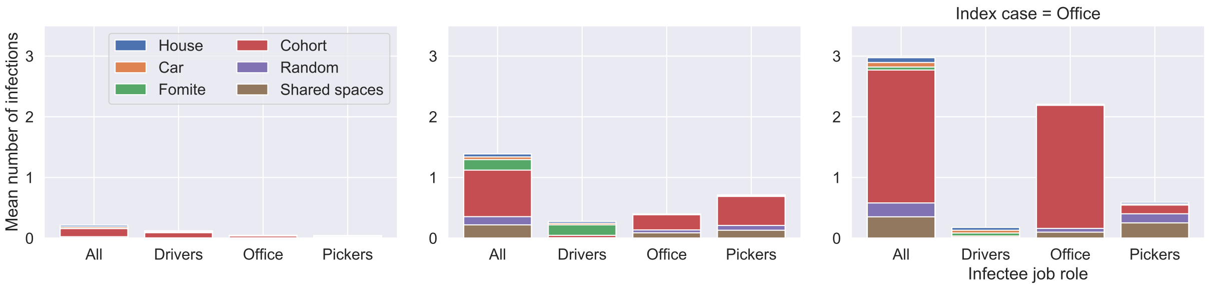


FIG. S4. Stacked bar charts of the mean number of simulated secondary infections resulting from a single index case in (a) a driver, (b) a picker, or (c) an office worker in the SPDD work setting. Each bar shows secondary infections in each group of staff broken down by transmission route, as recorded in table 2 in the main text. Note that the "shared spaces" contacts does not include contacts from sharing an office, these are counted as "cohort" interactions for office staff.

#### S3.1. BASELINE TRANSMISSION RATES BY JOB ROLE AND CONTACT TYPE

At baseline (assuming all default parameters in table 1 (main text), but high symptomatic isolation rate of $p_{\text{isol}} = 0.9$), our model predicts that the average number of cases the index case will infect in the parcel delivery workplace is $0.140 \pm 0.004$, $0.653 \pm 0.012$, and $2.24 \pm 0.03$ when the index-case is a driver, picker, or office worker respectively. In the large-items delivery work setting, it is predicted to be $1.398 \pm 0.016$, $0.982 \pm 0.015$, and $1.338 \pm 0.017$ when the index-case is a driver, picker, or office worker respectively. Note that the error bounds here are 1.96 standard errors of the mean, and do not account for parameter uncertainty (only stochasticity of the simulations).

Given the numbers of workers in each job role (see table 1 in the main text) this means that if the index case is selected at random we expect an R number of $\lesssim 1$ in the parcel work-setting, and $\approx 1$ in the large-item setting. Therefore, without any interventions our model predicts that these setting are very likely to see small outbreaks. Given the uncertainty in the underlying parameters, these baseline characteristics need to be considered as a "best guess" case. Bearing this in mind, there remains a great deal to be learned from quantifying the relative impact of various non-pharmaceutical interventions.

The average number of secondary cases in each job role and via each contact route is summarised for each workplace in figures S4 and S5.

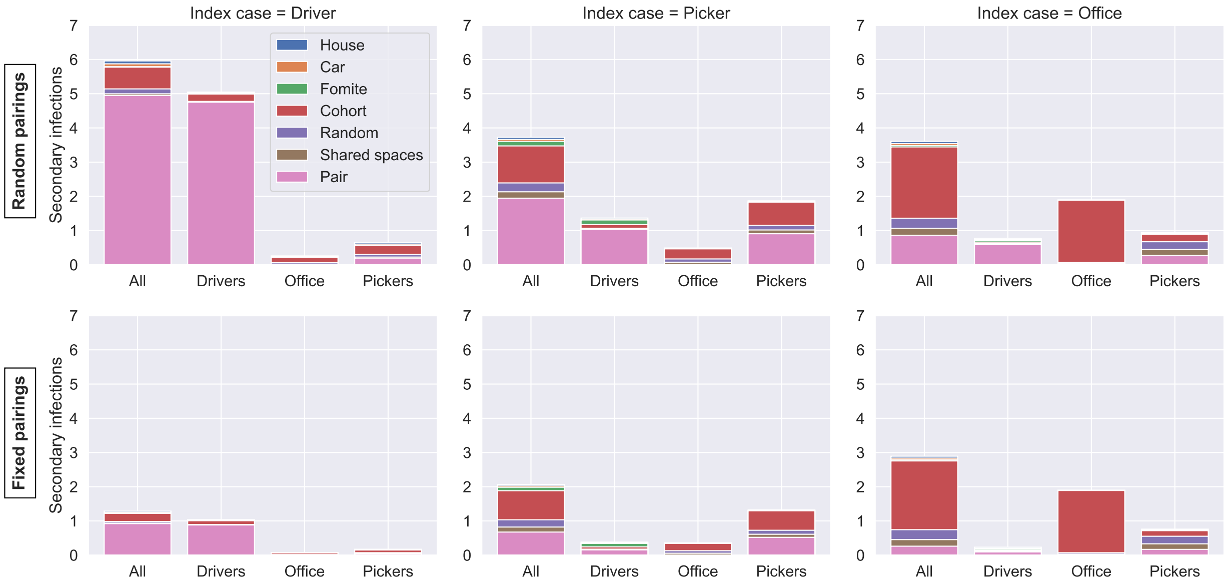


FIG. S5. Stacked bar charts of the mean number of simulated secondary infections resulting from a single index case in (a) a driver, (b) a picker, or (c) an office worker in the LIDD setting. Each bar shows secondary infections in each group of staff broken down by transmission route, as recorded in table 2 in the main text. Note that the "shared spaces" contacts does not include contacts from sharing an office, these are counted as "cohort" interactions for office staff.

#### S3.2. PARCEL DELIVERY: ROLE OF COHORT SIZE AND MIXING

Figure S6 shows the effect of increasing the number of work cohorts (and thereby reducing cohort size) in the parcel delivery work setting, as well as changing the rate at which staff switch between cohorts. Cohort-size only makes a large difference in the case where the index case is a member of office staff. This is because separating into different offices reduces overall rates of aerosol transmission. In the case of driver/picker cohorts, the underlying model assumptions mean that increasing the cohort size does not change the total amount of contact time (it is simply spread over more contacts). Therefore cohort size only makes a difference to outbreak probability when there is significant enough transmission via this route that saturation effects can occur. In our model this is the case for picker staff, where the effect is small, but not drivers.

Similarly, figure S6 also shows that increasing the rate at which employees move between cohorts ($f_c$) has very little effect on outbreak probability. Drivers are predicted to be much less likely to cause an outbreak due to their reduced time spent in the workplace.

Thus we predict that, for these settings, cohort size effects are most important when considering shared indoor spaces such as offices. In well ventilated spaces, where workers are generally spread out and only make contact intermittently, the dominant factor to consider is the total F2F contact time that each employee has with colleagues (not the number of distinct contacts). Note this ceases to be true for longer interactions where transmission risk is higher, as will be demonstrated in the following section.

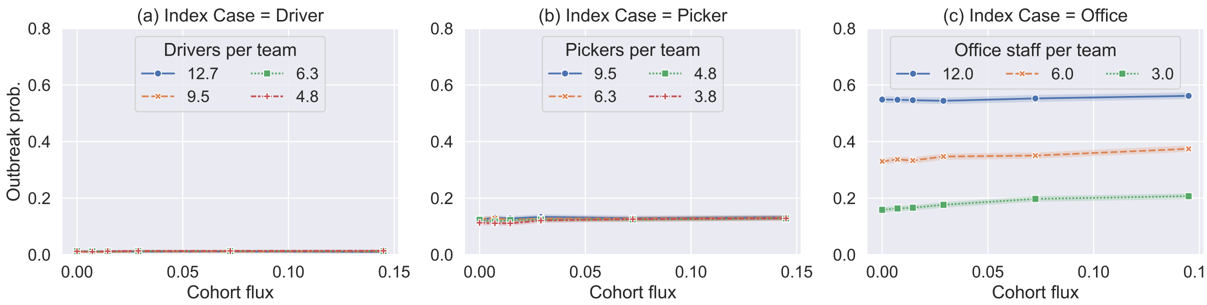


FIG. S6. Estimated probability of outbreak (defined as more than 3 secondary cases) resulting from a single index case plotted against the cohort flux $f_c$ in days$^{-1}$. Each marker shows the mean of 10,000 simulations of the SPDD workplace, with shaded error region estimated using a bootstrapping process [1]. Point-source outbreaks where the source case was (a) a driver, (b) a picker; (c) an office worker. Each line in each figure compares simulations with different numbers of teams used for that job role, shown as the number of workers per team on average. In each figure, the job roles not shown have the default team size and $p_{\mathrm{isol}} = 0.9$ is assumed.

#### S3.3. LARGE-ITEM DELIVERY: ROLE OF PAIR WORK AND LONGER CUSTOMER EXPOSURE

We now consider the case of workplaces that deliver large-items (e.g. furniture, white goods, etc.), such that drivers and pickers work in pairs to perform their tasks. This means they are in close-contact with one another for long periods of the day, increasing the risk of transmission. In figure S7 we consider 4 intervention scenarios:

1. No intervention: pairings are picked randomly at the start of each shift.

2. Fixed pairings: pairs are fixed and isolation of one partner automatically triggers isolation of the other.

3. Windows open: transmission rate between drivers sharing a cabin assumed to be the same as outside transmission rates, rather than inside.

4. Combination of scenarios 2 + 3.

Figure S7(a) shows the sizeable impact that the fixed pairings intervention has on the baseline dynamics. In particular, when the index case is a driver, the baseline dynamics are dominated by pair transmission (as shown in supplementary figure S5), and more-so for driver pairs due to the time spent in a shared cabin, which we assume to be higher risk than close-contact transmission in better-ventilated settings, such as the warehouse or on the doorstep. The randomly switching pairs mean that all drivers are connected by this transmission route, resulting in a larger number of secondary cases. Fixed pairings (scenario 2) close off these chains, meaning drivers can infect at most one person via this route, and that person then cannot infect anybody else via this route.

On the other hand, when the index case is a warehouse worker, figure S7(a) shows that changing from random to fixed pairings only has a relatively small effect on transmission. This indicates that, daily contact within picker-pairs are not predicted to be so high-risk to see a saturation effect. Note that this result is sensitive to the underlying parameterisation, and so has significant uncertainty associated with it.

Figure S7(b) compares the risk to customers related to a point-source outbreak with a driver index case in the large-item delivery workplace vs. the parcel workplace. It is clear that, for each individual customer there is increased risk (due to the longer contact time, chance of home entry needed). Nonetheless, the total number of customers likely to be infected is comparable in the two cases, owing to the large number of deliveries carried out by each driver in the parcel delivery setting. In both cases, even without interventions, the rate is predicted to be small ($0.144 \pm 0.008$ and $0.58 \pm 0.02$ customers infected per simulation in the parcel and large-item settings respectively, reducing to $0.124 \pm 0.008$ in the large-item workplace with 'FP + WO' interventions). Given a successful workplace outbreak (i.e. more than 5% of the workforce infected), this rate increases to $0.56 \pm 0.16$ and $1.06 \pm 0.03$ customer infections for the parcel and large-item delivery workplaces respectively, reducing to $0.45 \pm 0.05$ for the latter with 'FP + WO' interventions. Note that, in practice, customer infections were largely mitigated against via contactless delivery, which was implemented by the companies we consulted.

We conclude that pair-work can pose a high-risk of transmission in these settings, in which case fixed pairings in combination with pair isolation is an effective measure to break transmission chains and reduce the probability of a workplace outbreak. Generally, we predict that home delivery is unlikely to seed a significant number of new infections in the community, but it is likely that one or more customer infections will occur in the presence of a workplace outbreak. Thus, extra measures to protect customers (contactless delivery, mask wearing indoors etc.) will have likely made a meaningful impact when prevalence in the workplace was high.

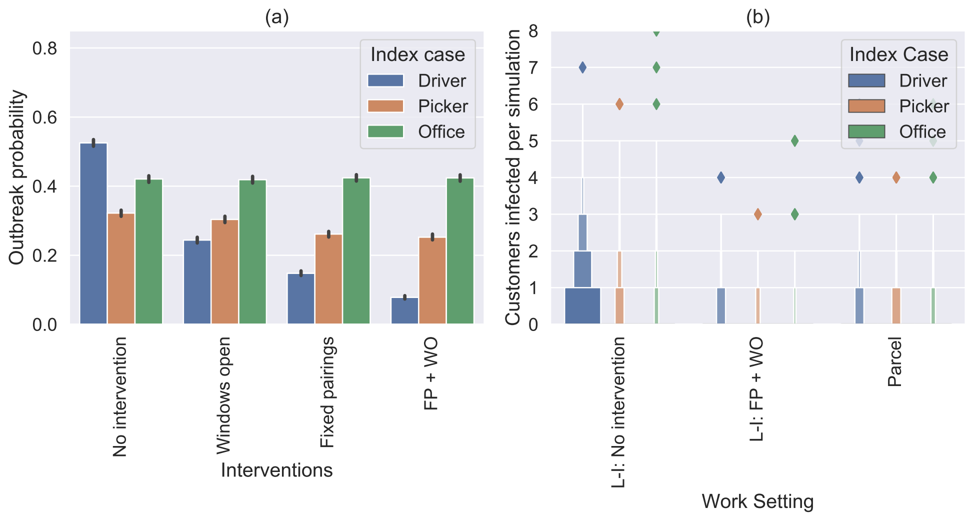


FIG. S7. (a) Simulated probability of an outbreak (defined as more than 2 secondary cases). Four scenarios are shown: no intervention (staff are randomly paired each day); driver pairs travel with window open (transmission rate constant reduced to 1/5 of original value in this setting); fixed pairs (people always work with the same partner); and both of these interventions simultaneously (fixed pairs and windows open). Each bar represents 10,000 simulations, error bars indicate uncertainty in the mean, estimated via a bootstrapping method [1]. (b) Boxen plots of the number of customers infected per point-source outbreak simulation in the LIDD setting with either no or both interventions and the parcel delivery setting with default parameters.

#### S3.4. EFFECTS OF PRESENTEEISM

In this model we define presenteeism as working with symptoms of COVID-19 (as we do not model other sickness absences explicitly). On average, 50% of the employees in our simulations develop symptoms relevant for isolation, and if they isolate then they do not attend work for the following 10 days (but household transmission between employees is assumed unaffected). Note that we do not model the effects of illness severity (e.g. prolonged symptoms after recovery, hospital stays, or even death).

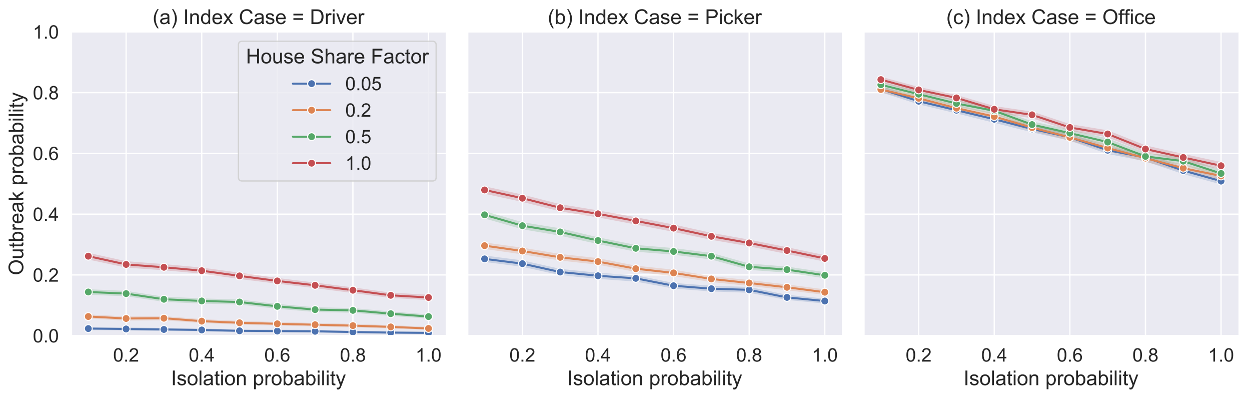


FIG. S8. Dependence of simulated outbreak probability on the self-isolation adherence probability $p_{\text{isol}}$ in the model SPDD work setting. The different curves show the effect of increasing the house-sharing factor $H$ as labelled.

Figure S8 shows the effect of increasing self-isolation rates among symptomatic individuals in both work settings, for various values of $H$, the house-sharing factor. Outbreak probability reduces linearly with isolation adherence, and the proportional effect is larger when house-sharing is rare.

Figure S9 shows the impact of $p_{\text{isol}}$ in the large-item delivery setting on secondary cases (rather than outbreak probability). The impacts are similar in this workplace to the parcel setting.

Focusing on when the index case is a driver: the fixed pairings policy has a large impact on the mean number of secondary cases. Increasing adherence to symptomatic isolation has a similar relative effect in both cases ($\sim$50% reduction in number of cases increasing $P_{\text{isol}}$ from 0.1 to 1.0 either with or without the fixed pairs policy). Comparing the transmission dynamics in these two cases we see that the increased rates of isolation are actually impacting different contact routes to different extents. When there is no fixed pairings, improved isolation adherence impacts infection via all contact routes by a similar amount proportionally ($\sim$50% reduction in number of cases increasing $P_{\text{isol}}$ from 0.1 to 1.0), see figure S10(a). In the case of fixed pairs interventions, the increase in isolation adherence more significantly reduces ($>$ 50% reduction) infections via all contact routes except the fixed pairings them-

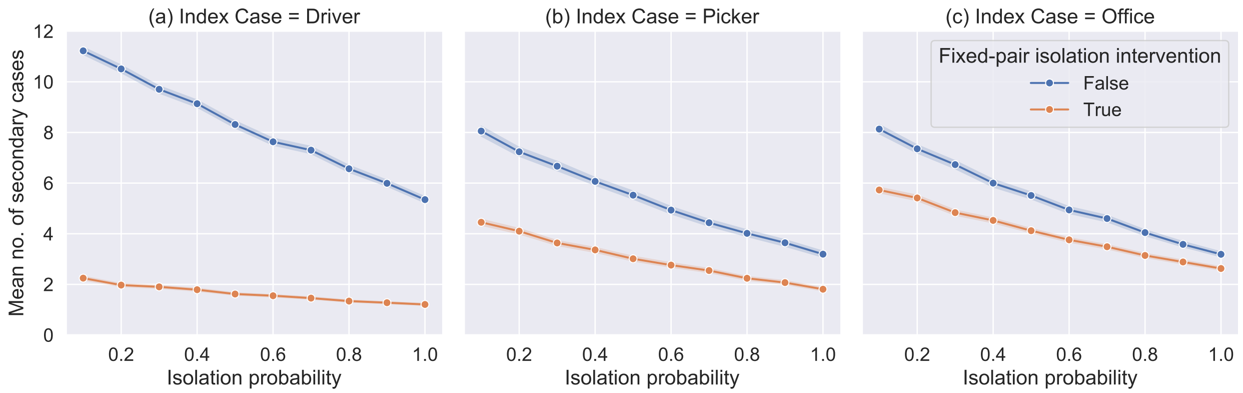


FIG. S9. Dependence of mean number of simulated secondary cases from a single index case on the self-isolation adherence probability $p_{\mathrm{isol}}$ in the model LIDD work setting. The different curves show the effect of adding a fixed-pairs isolation intervention, as described in the previous section.

selves ($<$ 50% reduction), see figure S10(b). This is because, in this case when a driver index case develops symptoms and isolates, they have likely already infected their (fixed) partner through daily close contact, whereas the isolation period stops them infecting other colleagues via the other contact routes. Moreover, when they isolate, their work partner also isolates (usually much earlier than they would develop symptoms) removing the vast majority of their potential onward workplace infection, which otherwise would not be via the 'pair' route anyway, because their work partner was the index case, and cannot be reinfected.

When the index case is not a driver, we see from figures S9(b) and (c) that the fixed pairings intervention has a more modest effect on overall infection counts as the pair infection route is less prominent in these transmission chains.

To conclude, adherence to isolation measures is predicted to make a substantial difference to transmission dynamics. The size of the impact is not obvious when considering the case of pair isolation policies in the large-item workplace, but nonetheless the measures were complimentary, greatly reducing the probability of transmission beyond the affected pair when the index case was a driver. We saw a less-strong effect for household isolation in our model even when a large fraction of workers share accommodation, this is because we assumed that this measure does not prevent household transmission, which was the dominant transmission mode in this scenario. This stresses the importance of messaging

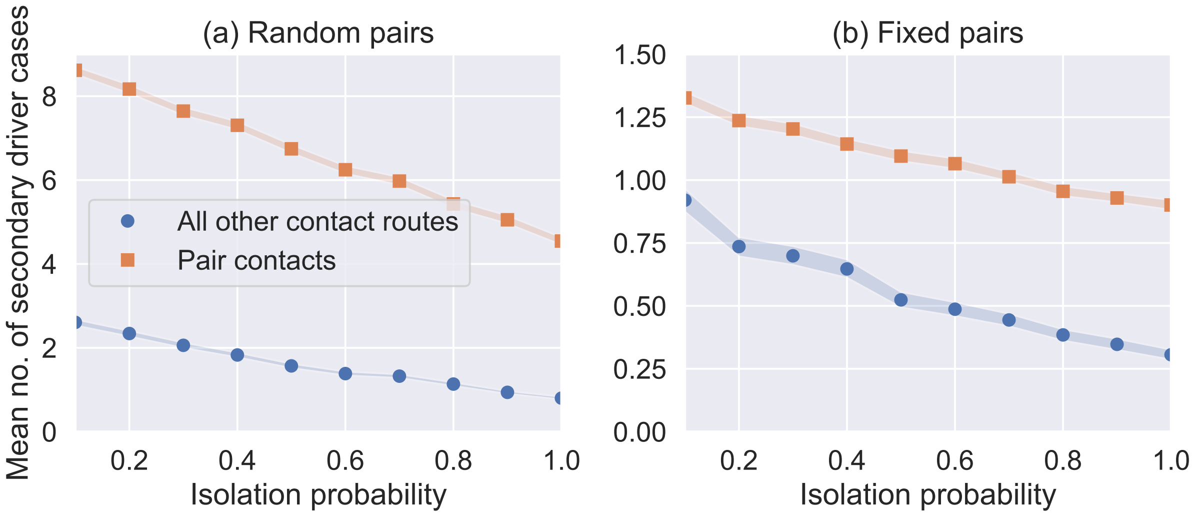


FIG. S10. Mean number of infected drivers per simulation with a single driver index casein the LIDD setting, plotted against symptomatic isolation probability $p_{\mathrm{isol}}$. The infections are broken down by those cased by close contact pair work, and all other contact routes. (a) The case with no fixed pairing intervention so pairs switch randomly each day. (b) The case with fixed pairings a pair isolation policy. Dots show the mean number of infections while shading shows 95% confidence in the mean calculated via bootstrapping methods.

around measures to reduce within-household transmission as well isolation measures.

[1] Michael L Waskom. Seaborn: Statistical data visualization. *J. Open Source Softw.*, 6(60):3021, 2021. doi:10.21105/joss.03021.
