## Supplementary material for "Modelling the impact of non-pharmaceutical interventions on workplace transmission of SARS-CoV-2 in the home-delivery sector": All supplementary files: SupplementaryTextS4.pdf

### Supplementary Text S4 for "Modelling the impact of non-pharmaceutical interventions on workplace transmission of SARS-CoV-2 in the home-delivery sector": Sensitivity Analysis

Carl A. Whitfield,[1,2,3] Martie van Tongeren,[4,3] Yang Han,[1] Hua Wei,[4,3] Sarah Daniels,[4,3] Martyn Regan,[5,4,3] David W. Denning,[2,3] Arpana Verma,[4,3] Lorenzo Pellis,[1] University of Manchester COVID-19 Modelling Group,[1] and Ian Hall[1,6,3]

[1]*Department of Mathematics, University of Manchester, Manchester, England*

[2]*Division of Infection, Immunity & Respiratory Medicine, School of Biological Sciences, University of Manchester, Manchester, England*

[3]*Manchester Academic Health Science Centre, University of Manchester, Manchester, England*

[4]*Division of Population Health, Health Services Research & Primary Care, School of Health Sciences, University of Manchester, Manchester, England*

[5]*National COVID-19 Response Centre, UK Health Security Agency, London, England*

[6]*Public Health Advice, Guidance and Expertise, UK Health Security Agency, London, England*

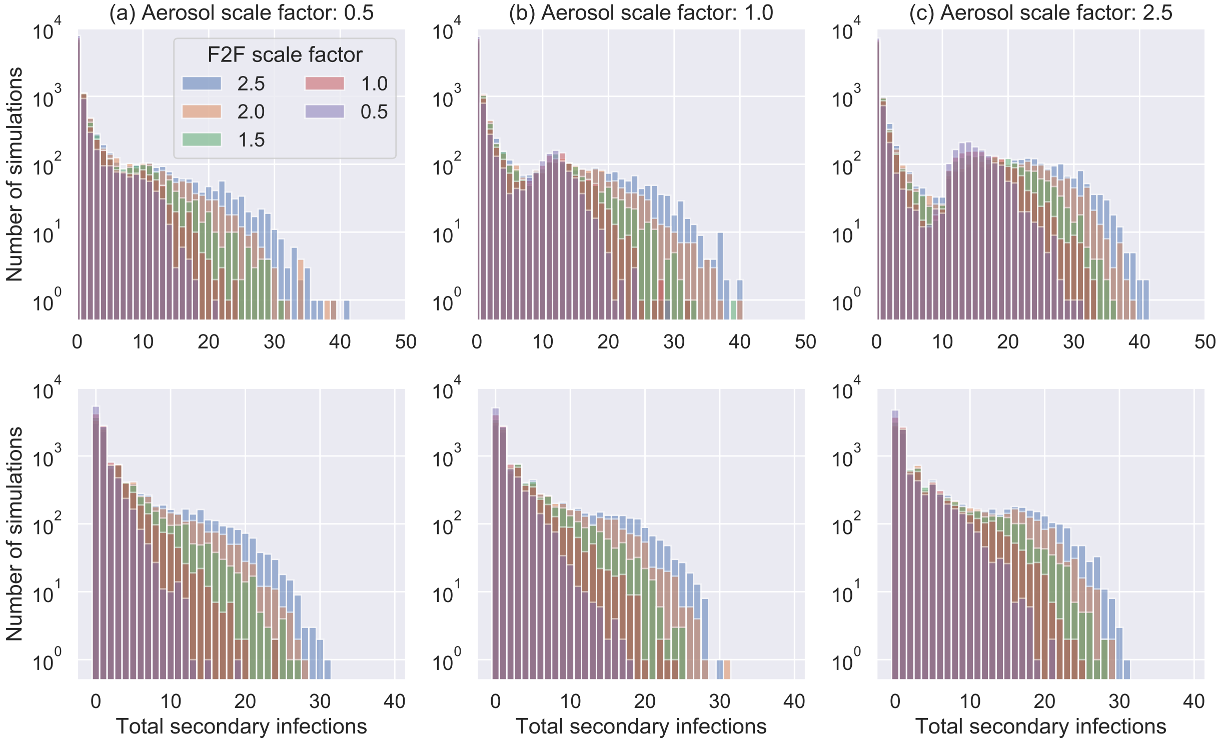


FIG. S11. Histograms of secondary cases resulting from a single index case in the two work settings simulated for different rates of F2F and aerosol transmission. The top row shows the parcel work setting, while the bottom row is the large-item setting. For each set of simulations, the transmission rate for F2F contacts is multiplied by “F2F scale factor”, and the transmission rate for aerosol contacts is multiplied by “Aerosol scale factor”. Note that for the large-item workplace we assume that the fixed-pair isolation intervention is applied and in both cases $p_{\mathrm{isol}} = 0.9$. We also assume that the index case is selected randomly.

#### S4.1. TRANSMISSION PARAMETERS

The baseline transmission rate for F2F contacts is derived from multiple sources, but these are largely observational studies and involve a number of assumptions. It is therefore useful to observe how some of the key results are affected if the transmission rate is higher (e.g. due to more transmissible variants, or due to error in the transmission rate used). Figure S11 shows how the histogram of secondary case numbers changes as the transmission rate $\beta_{F2F}$ is increased (note we assume that shared-space transmission also increases proportionally). Assuming a random index case, we see a transition from an exponential-type distribution to a multi-modal distribution, as the workplace R-number exceeds 1 and most introductions result in several secondary cases. In these cases, NPIs can have a much more significant impact, as they can push the workplace R-value back to below 1.

Second, the fomite transmission parameter is an unknown in the model, so it is important to see for what values it must take for fomite transmission to be significant. In figure S12 we see that increasing the fomite transmission by a factor of 10 makes a relatively minor difference to the outcome of point-source outbreaks originating with a picker (it has a much smaller effect for other index cases). However, the change is substantial when increasing by another factor of 10, suggesting that, if this were the case, it would be a dominant mode of transmission and a significant contributor to overall workplace transmission. Note that, even though the effect is much less significant in the large-items workplace, the $\beta_{FOM}$ value might actually be larger in this environment, since these items require more force to move (which can affect surface deposition rates) and may require more overall close-contact with the item.

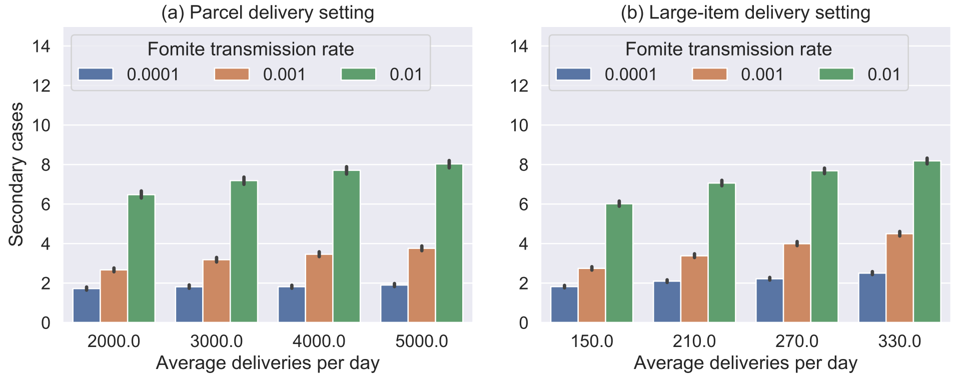


FIG. S12. The mean number of secondary cases resulting from a single index case in the two workplace types plotted for 3 values of $\beta_{\mathrm{FOM}}$ at varying levels of demand for deliveries (x-axis). Note that for the large-item workplace we assume that the fixed-pair isolation intervention is applied and in both cases $p_{\mathrm{isol}} = 0.9$.

#### S4.2. NUMBER OF EMPLOYEES

The influence of overall workplace size is largely controlled by how we assume the contact networks scale in warehouses with fewer/more staff. In order to test this we use simulate workplace sizes corresponding to the upper and lower quartiles of the datasets in Supplementary Text S1. We scale the workplace size by factor $0.5 \leq r \leq 2$ in the parcel setting and $0.5 \leq r \leq 3$ in the pairs setting. We assume that the parameters scale as follow

$$\{N_D, N_L, N_O\} \rightarrow \{\lfloor rN_D \rceil, \lfloor rN_L \rceil, \lfloor rN_O \rceil\} \tag{1}$$

$$\{T_D, T_L, T_O\} \rightarrow \{\lfloor rT_D \rceil, \lfloor rT_L \rceil, \lfloor rT_O \rceil\} \tag{2}$$

while the definition of $p_c$ remains the same (and so decreases with workplace size).

Figure S13 shows the results of point-source outbreak simulations in the two settings across the range of feasible workplace size scalings. We see that the number of secondary cases resulting from the outbreak increases with workplace size, although this is, in part, explained by the increase in office cohort size (the sharp reduction occurs when the number of teams is increased). This is because the number of teams are chosen relative to the number of employees (to keep the number per team as consistent as possible). However, these changes in team numbers occur at discrete thresholds, and as shown in figure S6. This shows that some of the effects of changing workplace size are simply due to office sizes increasing (the effect shown in figure S6), but that this does not completely account for all of the increase in infections with size.

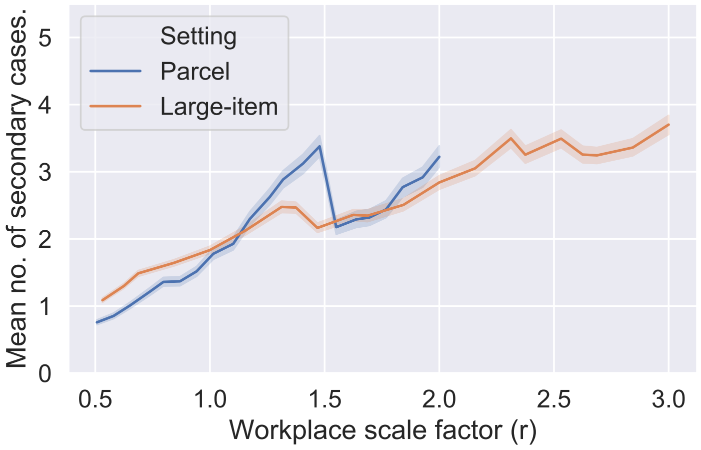


FIG. S13. The mean number of secondary cases resulting from a point-source outbreak in the two workplace types plotted against workplace scale factor. Note that for the large-item workplace we assume that the fixed-pair isolation intervention is applied and in both cases $p_{\text{isol}} = 0.9$. We assume the index case is selected at random.

It is worth noting though that, with greater mixing rates than simulated here, larger workplaces could pose a larger risk of outbreak because of the presence of a larger susceptible population.

#### S4.3. MIXING RATES

We vary the amount of mixing in the workplace by changing the parameter $p_c$. As $p_c$ increases, not only do more contacts occur but also contacts between different job roles become more frequent. The parameter $\rho_D$ remains fixed at 0.1, so drivers still have less contact than the other job roles. Figure S14 shows how the number of secondary cases in a point-source outbreak increases as $p_c$ is increased for each workplace.

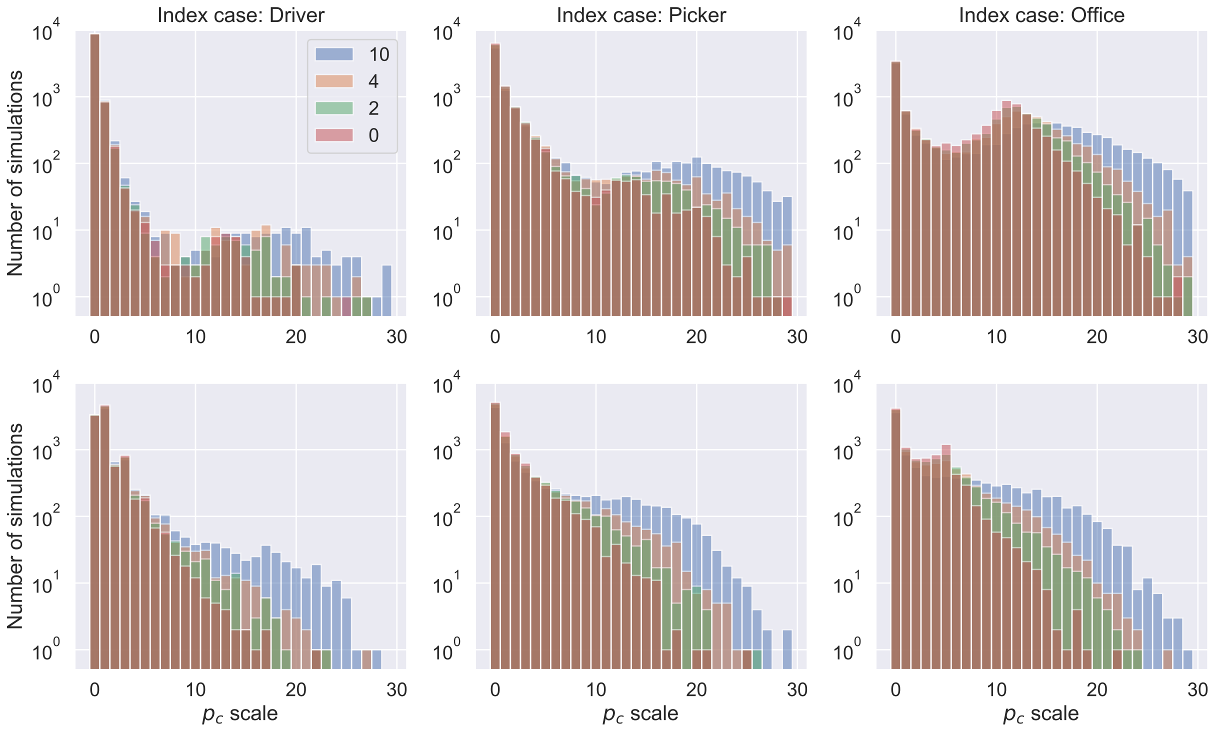


FIG. S14. Histograms of the number of secondary cases resulting from a single index case in the two workplace types plotted for different scalings of $p_c(N_D + N_L + N_O)$. The top row shows the parcel delivery setting, while the bottom row is large-item setting, and each column is for the index-case labelled. Note that for the large-item workplace we assume that the fixed-pair isolation intervention is applied and in both cases $p_{\text{isol}} = 0.9$. Note also the logarithmic scale.
