## Supplementary figures and images for "Modelling the impact of non-pharmaceutical interventions on workplace transmission of SARS-CoV-2 in the home-delivery sector"

### figureS1.pdf

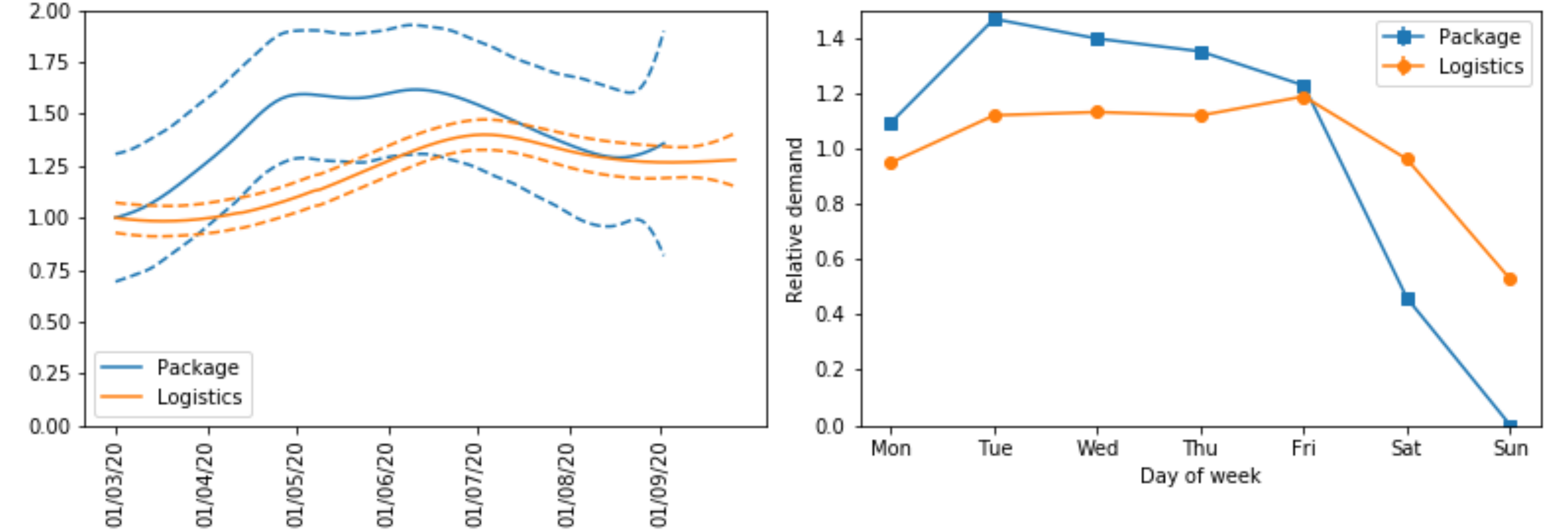

### figureS2.pdf

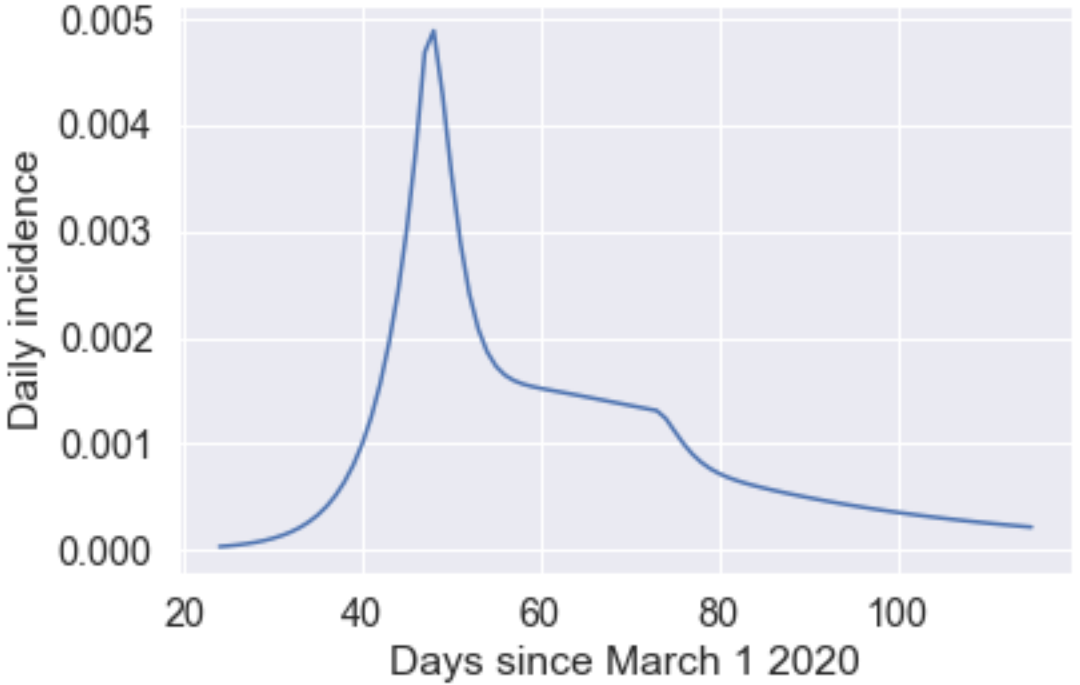

### figureS3.pdf

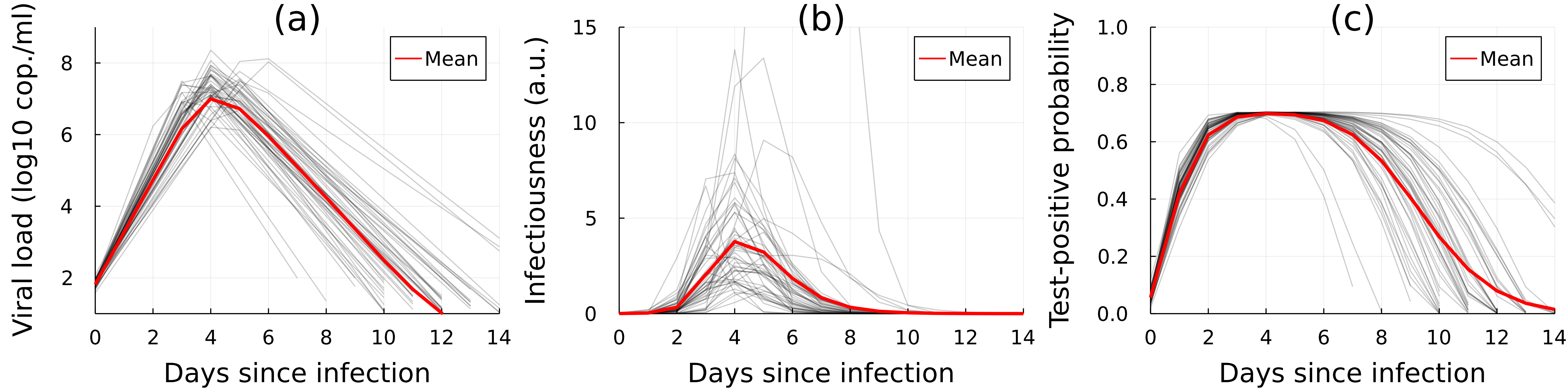

### figureS4.pdf

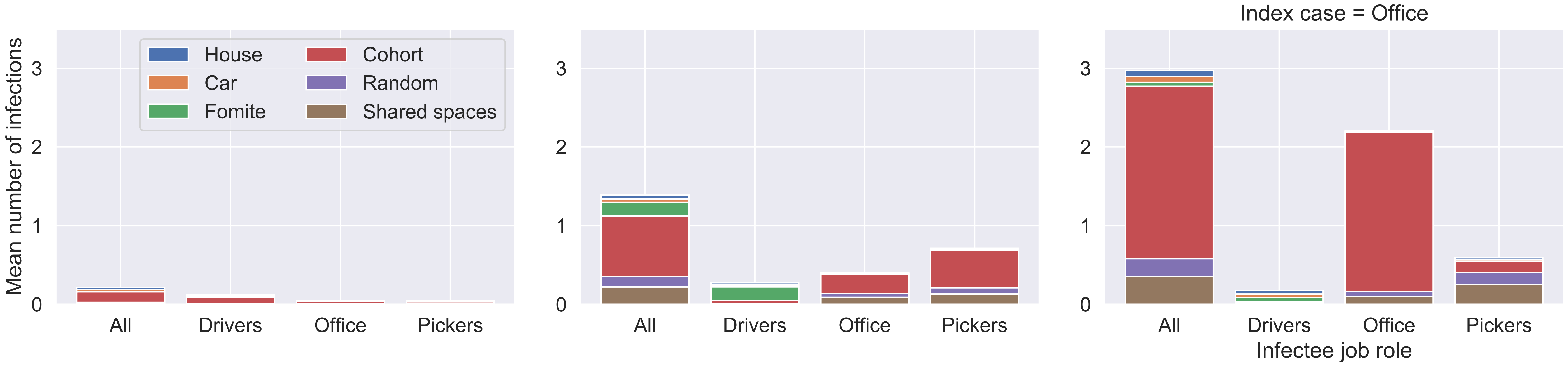

### figureS5.pdf

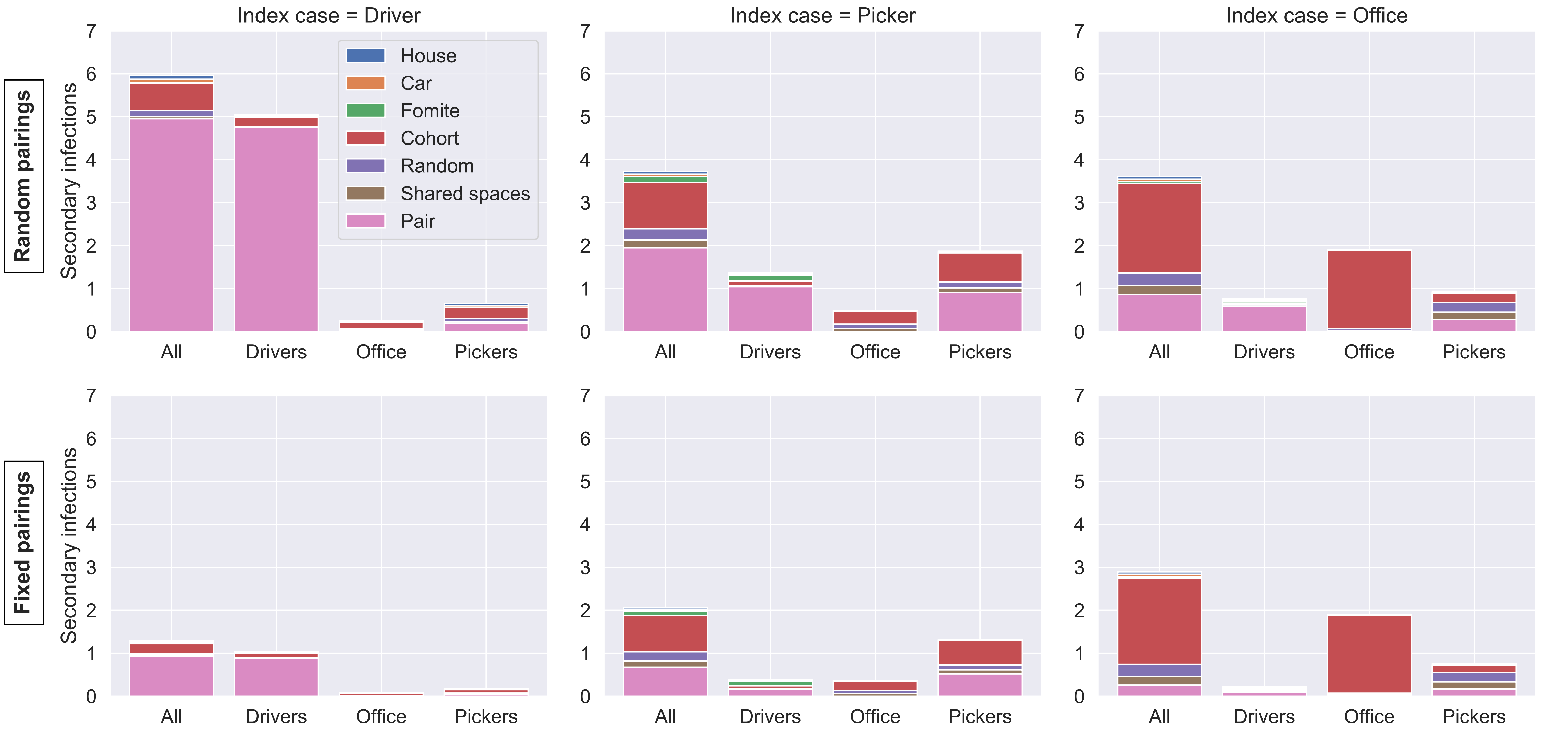

### figureS6.pdf

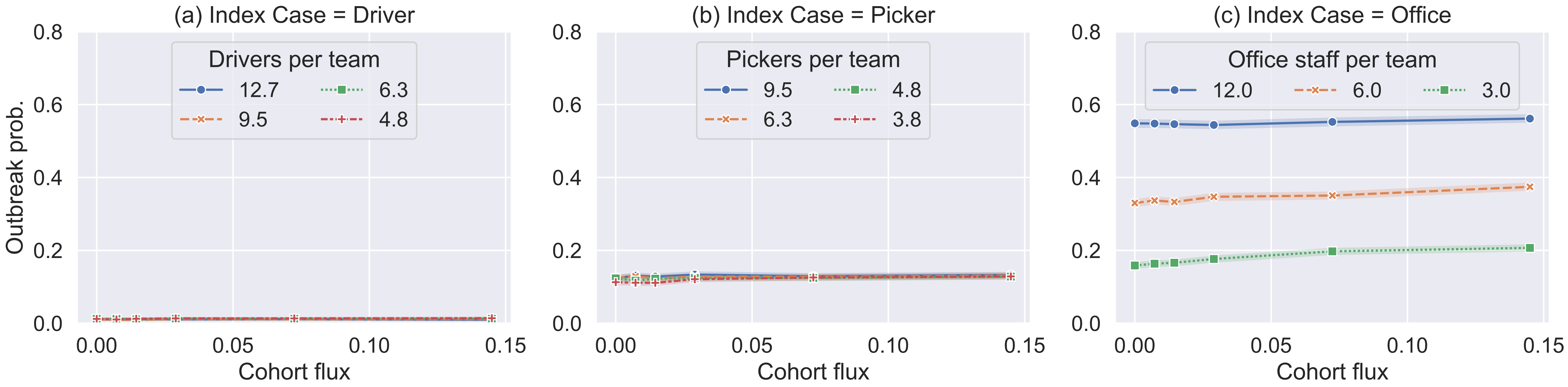

### figureS7.pdf

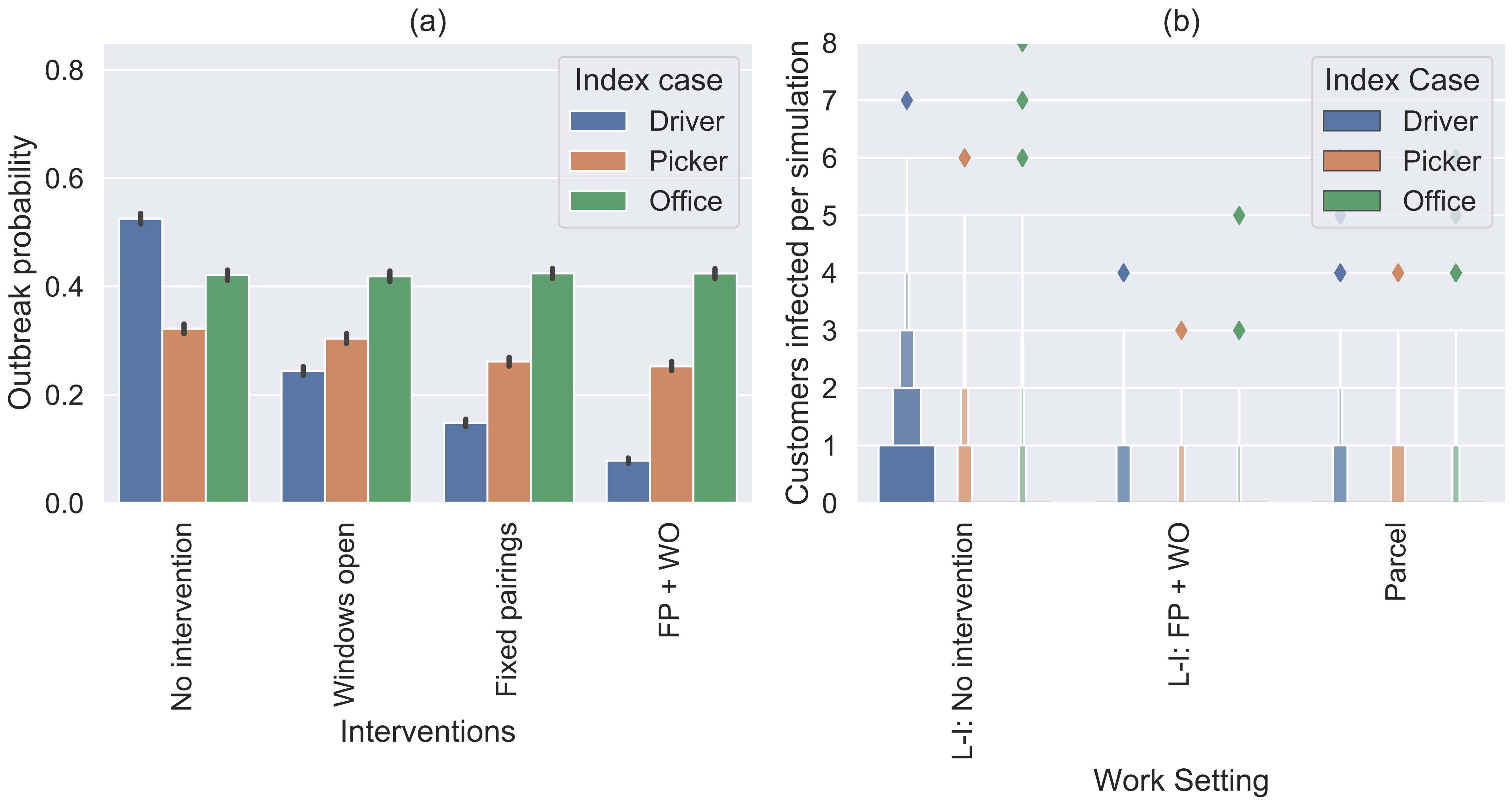

### figureS8.pdf

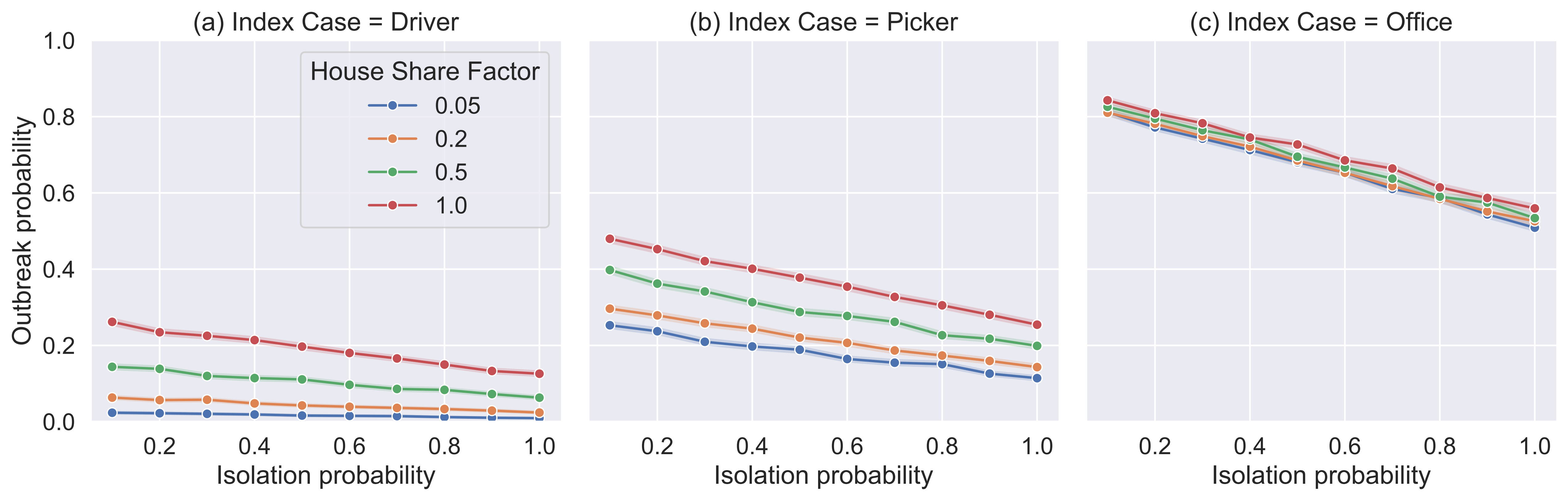

### figureS9.pdf

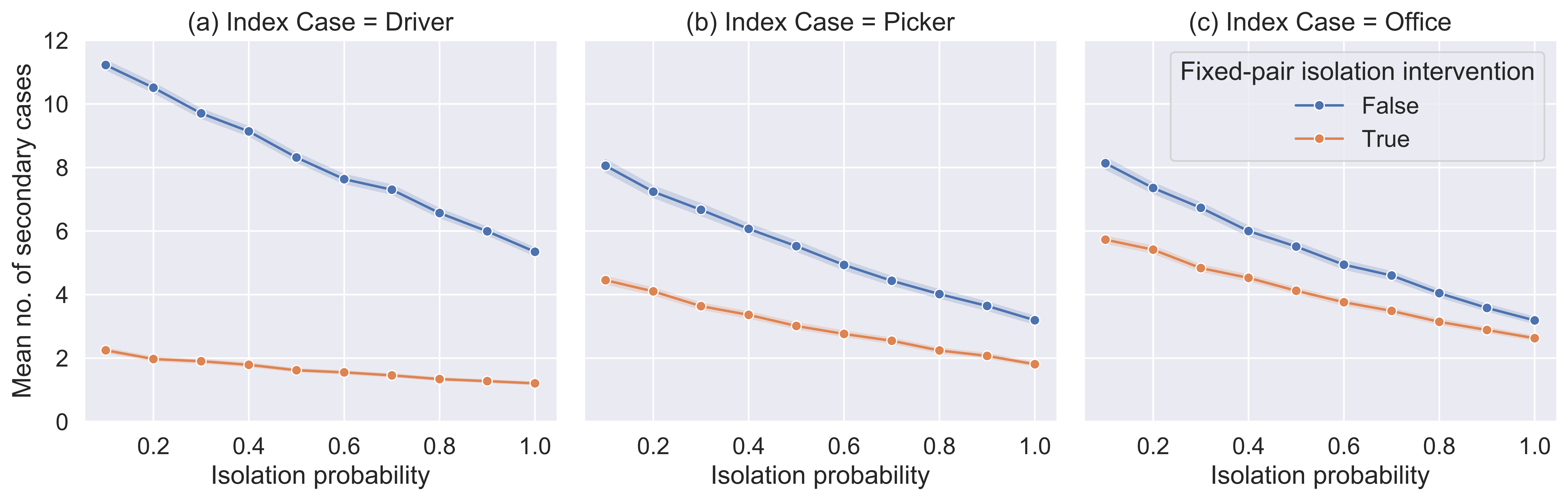

### figureS10.pdf

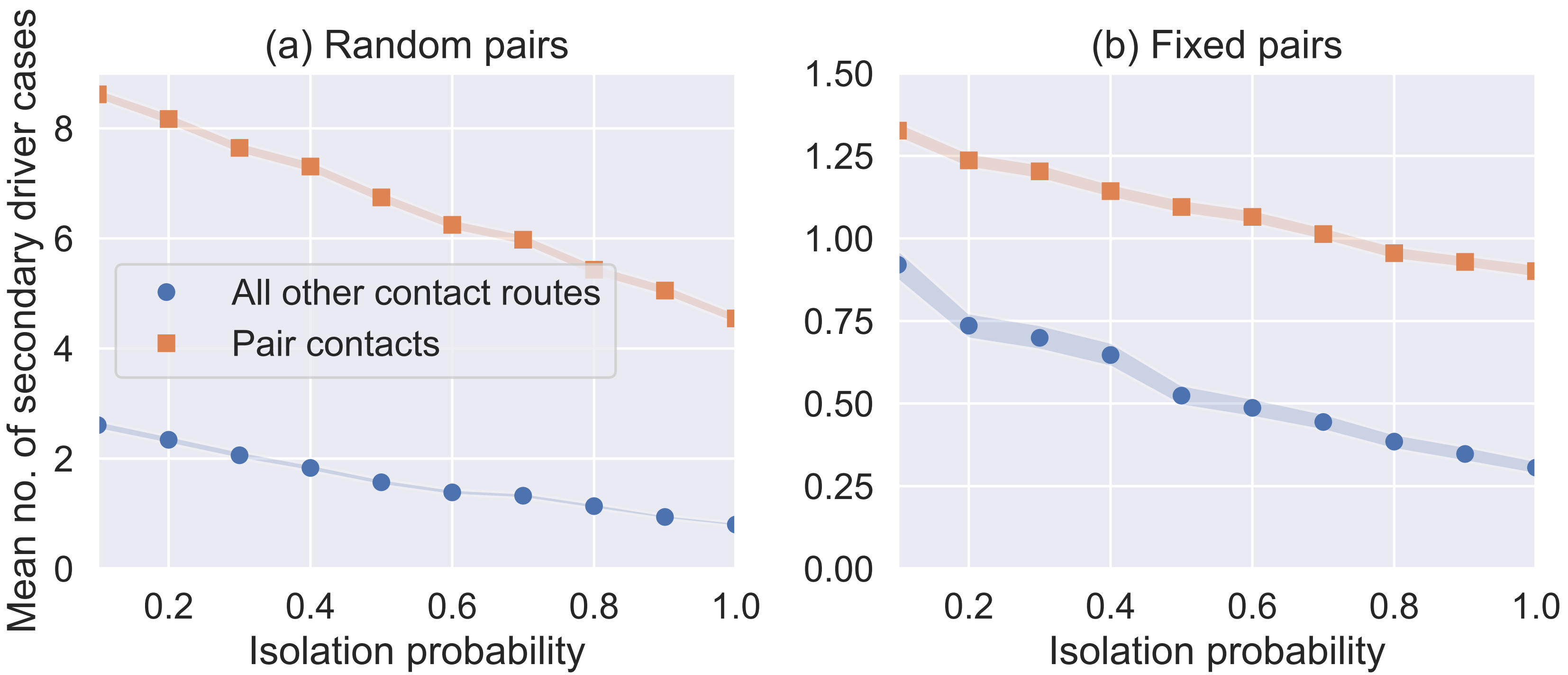

### figureS11.pdf

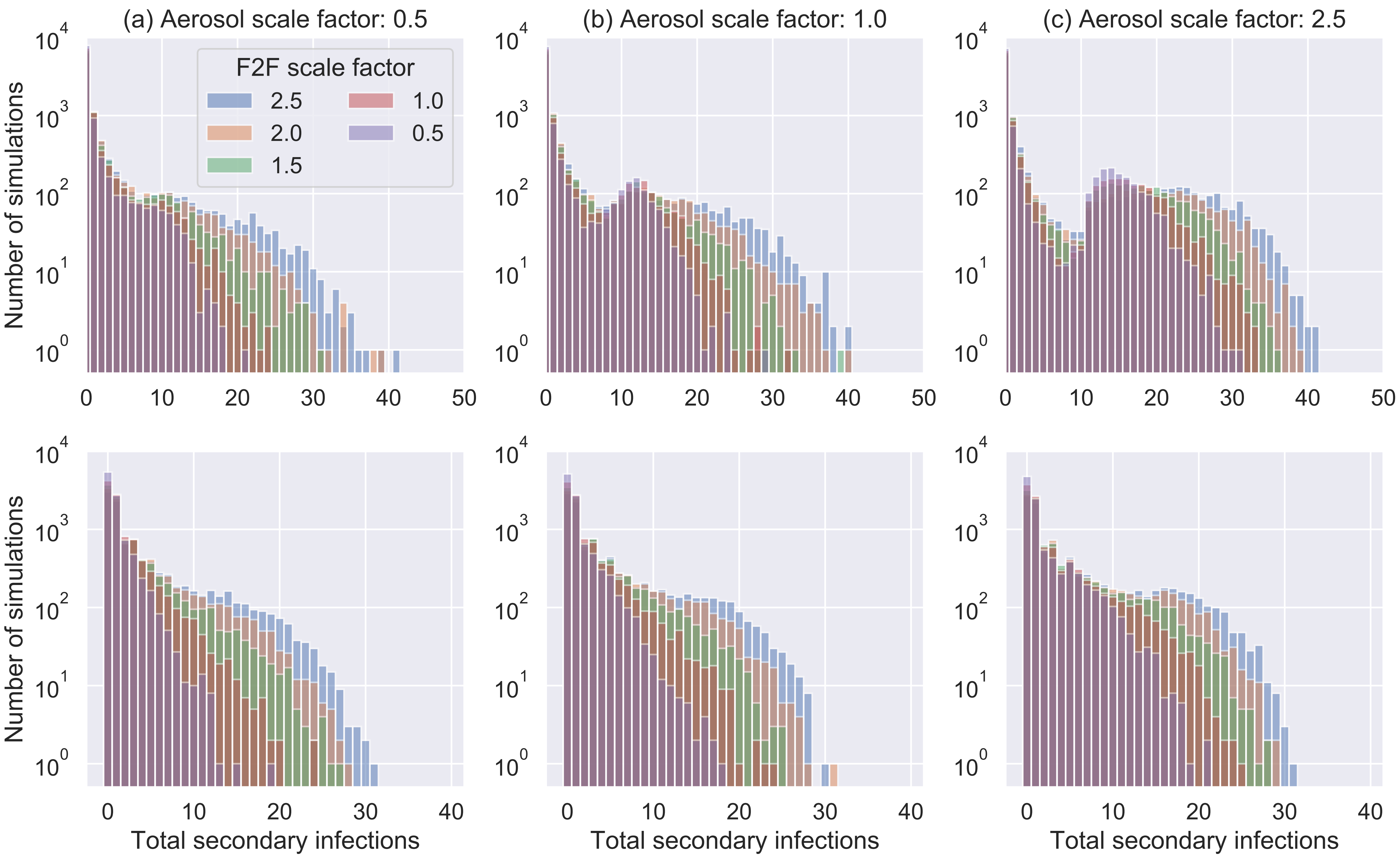

### figureS12.pdf

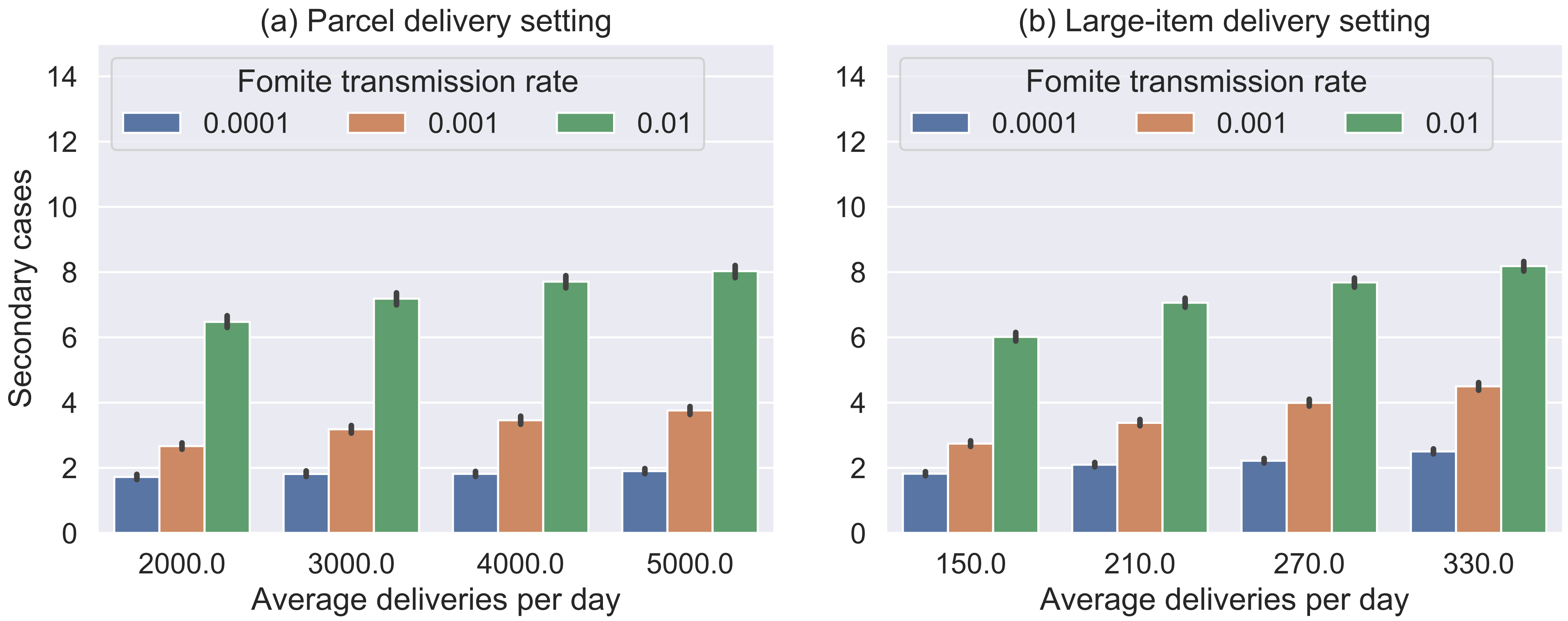

### figureS13.pdf

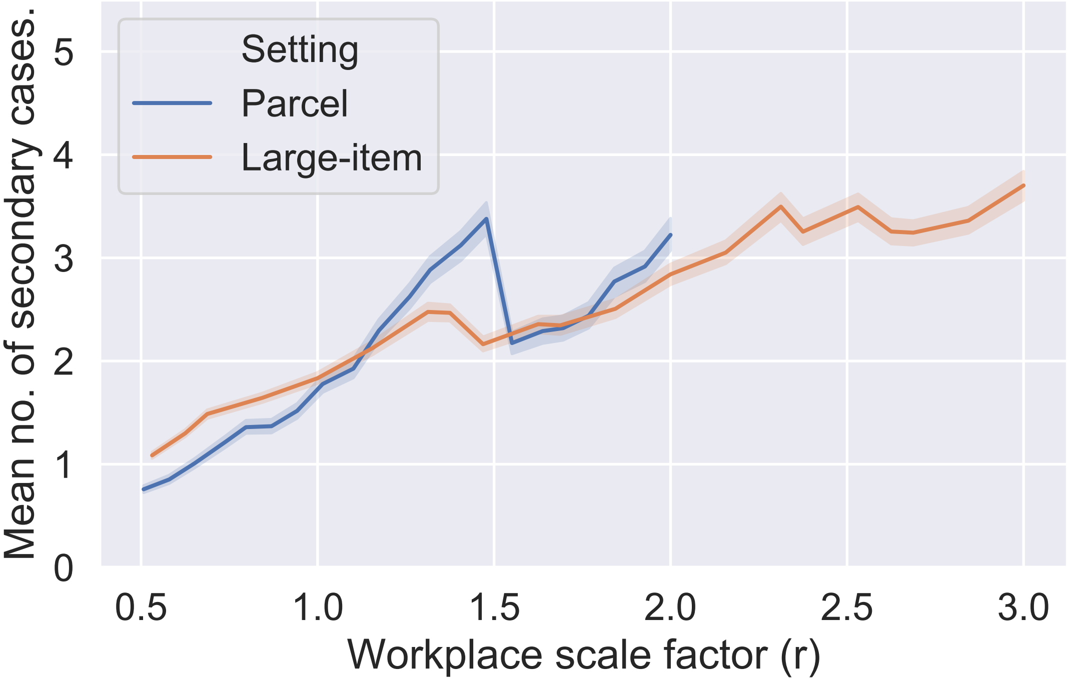

### figureS14.pdf

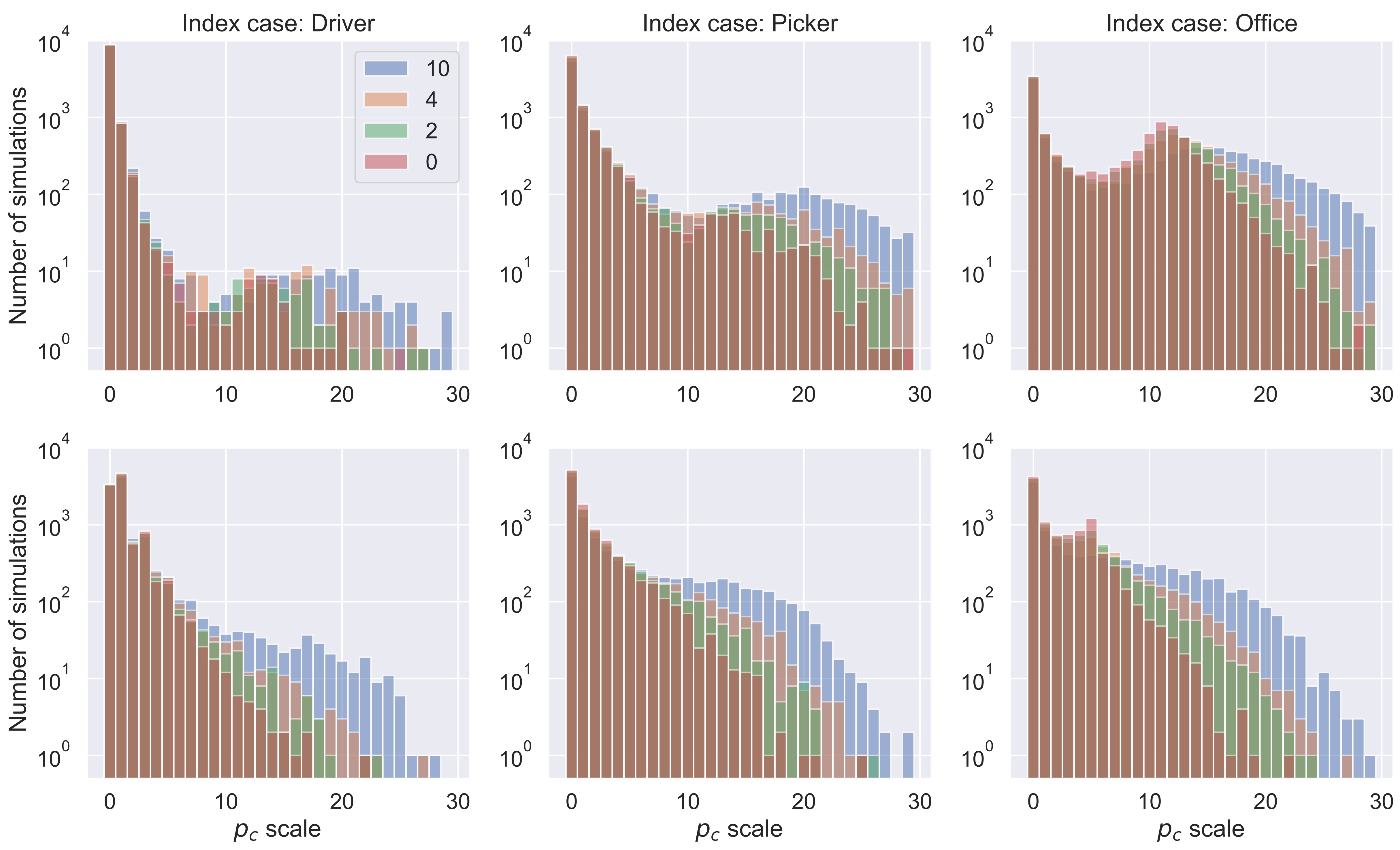
